# Changes in COVID-19-related mortality across key demographic and clinical subgroups: an observational cohort study using the OpenSAFELY platform on 18 million adults in England

**DOI:** 10.1101/2022.07.30.22278161

**Authors:** The OpenSAFELY Collaborative, Linda Nab, Edward PK Parker, Colm D Andrews, William J Hulme, Louis Fisher, Jessica Morley, Amir Mehrkar, Brian MacKenna, Peter Inglesby, Caroline E Morton, Sebastian CJ Bacon, George Hickman, David Evans, Tom Ward, Rebecca M Smith, Simon Davey, Iain Dillingham, Steven Maude, Ben FC Butler-Cole, Thomas O’Dwyer, Catherine L Stables, Lucy Bridges, Christopher Bates, Jonathan Cockburn, John Parry, Frank Hester, Sam Harper, Bang Zheng, Elizabeth J Williamson, Rosalind M Eggo, Stephen Evans, Ben Goldacre, Laurie A Tomlinson, Alex J Walker

## Abstract

**Objectives:** To quantify in absolute and relative terms how population-level COVID-19 death rates have changed in demographic and clinical subgroups.

**Design:** Retrospective cohort study on behalf of NHS England.

**Setting:** Linked primary care and death registry data from the OpenSAFELY-TPP platform, covering the first three pandemic waves in England (wave 1: March 23 to May 30, 2020; wave 2: September 7, 2020 to April 24, 2021; and wave 3, delta: May 28 to December 14, 2021).

**Participants:** In total, 18.7, 18.8, and 18.7 million adults were included for waves 1, 2, and 3 respectively.

**Main outcome measures:** COVID-19-related mortality based on linked death registry records.

**Results:** The crude absolute COVID-19-related death rate per 1,000 person-years decreased from 4.48 in wave 1 (95%CI 4.41;4.55), to 2.70 in wave 2 (95%CI 2.67;2.73), to 0.64 in wave 3 (95%CI 0.63;0.66). The absolute death rate decreased by 90% between waves 1 and 3 in patients aged 80+, but by only 20% in patients aged 18-39. This higher proportional reduction in age- and sex-standardised death rates was also seen for other groups, such as neurological disease, learning disability and severe mental illness. Conversely, standardised death rates in transplant recipients stayed constant across successive waves at 10 per 1,000 person-years. There was also only a small decrease in death rates between waves in people with kidney disease, haematological malignancies or conditions associated with immunosuppression. Consequently, the relative hazard of COVID-19-related death decreased over time for some variables (e.g. age), remained similar for some (e.g. sex, ethnicity), and increased for others (e.g. transplant).

**Conclusions:** COVID-19 death rates decreased over the first three pandemic waves. An especially large decrease was seen in older age groups and people with neurological disease, learning disability or severe mental illness. Some demographic inequalities in death rates persisted over time. Groups more likely to experience impaired vaccine effectiveness did not see the same benefit in COVID-19 mortality reduction.

## Background

COVID-19 has been shown to disproportionately affect subgroups of the population depending on their demographic and clinical profile. Previous studies in the UK have reported higher COVID-19-related mortality in groups of older age, male sex, non-White ethnicity, public facing occupations (e.g., social care workers), multigenerational living, social deprivation, learning disability, and clinical comorbidities such as obesity and kidney disease, among others [1]–[6].

Much of what is known regarding population subgroups at greater risk of COVID-19-related mortality comes from the early stages of the pandemic. During the third pandemic wave in the UK, which was dominated by the Delta variant, overall monthly COVID-19 mortality rates were considerably lower than the first and second pandemic waves [7]. This reduction may be attributed in part to the widespread implementation of COVID-19 vaccines among high-risk groups [8], as well as improvements in clinical management (e.g. timely administration of antivirals [9] or corticosteroids [10]) and non-pharmaceutical interventions [11] (e.g., physical distancing and shielding).

Despite these improvements, clinical and demographic inequalities in mortality burden may persist. For example, in a study of post-immunisation infection among individuals who had completed a 2-dose series of COVID-19 vaccines, population subgroups experiencing higher rates of COVID-19-related death – including individuals with kidney disease and malignancies – were similar to those seen at the start of the pandemic [12].

We therefore set out to quantify in absolute and relative terms how population-level COVID-19-related death rates have changed in demographic and clinical subgroups over the course of the pandemic. Here, we report on trends in COVID-19 mortality across clinical and demographic subgroups of the population between February 2020 and December 2021.

## Methods

### Data Source

Primary care records managed by the GP software provider TPP were accessed through the OpenSAFELY platform, where all data were linked, stored and analysed securely (https://opensafely.org/). Data include pseudonymised data such as coded diagnoses, medications and physiological parameters. No free text data are included. All code is shared openly for review and re-use under MIT open licence (https://github.com/opensafely/covid_mortality_over_time). Detailed pseudonymised patient data is potentially re-identifiable and therefore not shared.

### Study population

From 23 March 2020 - 14 December 2021, three cohorts were extracted covering the first three pandemic waves in England (wave 1: March 23 to May 30, 2020; wave 2: September 7, 2020 to April 24, 2021; and wave 3, delta: May 28 to December 14, 2021). Start and end dates of the first two pandemic waves were determined based on estimates published by the Office for National Statistics [13]. The start date of the third pandemic wave (May 28, 2021) was determined based on the day that reported reproduction was above 1 (1.0-1.1) and growth rates in England were positive (0-3) [13]. The end date (14 December 2021) was chosen as Omicron became nationally dominant after this date [14]. The three extracted cohorts consisted of individuals aged between 18 years and 110 years registered with a TPP practice on the first day of the wave with at least three months of continuous GP registration prior to this date, to ensure that baseline data could be adequately captured. People were excluded if they had missing data for sex or demographic data (i.e. the Sustainability and Transformation Partnership region [STP, an NHS administrative region], or index of multiple deprivation [IMD]).

### Study measures

#### Outcomes

The outcome of interest was COVID-19-related mortality based on linked death registry records from the Office for National Statistics. COVID-19 deaths were defined as having an underlying or contributory cause of death listed as COVID-19 (ICD-10 codes U07.1 or U07.2).

#### Covariates

Covariates considered in the analysis included health conditions listed in UK guidance on higher risk groups [15], other common conditions that may cause immunodeficiency inherently or through medication, and other postulated risk factors for severe outcomes among COVID-19 cases. We included age (grouped as 18-39, 40-49, 50-59, 60-69, 70-79 and ≥80 years for descriptive analysis), sex, ethnicity (White, Mixed, Asian, Black, Other, Unknown), body mass index (BMI; categorised as not obese, class I [BMI 30-34.9kg/m2], II [35-39.9kg/m2], III [≥40kg/m2]), smoking status (never, former, current), and index of multiple deprivation quintile (derived from the patient’s postcode at lower super output area level). We also considered the following comorbidities: high blood pressure (systolic blood pressure ≥140 or diastolic blood pressure ≥90) or diagnosed hypertension, chronic respiratory diseases other than asthma, asthma (categorised as with or without recent use of oral steroids), chronic cardiac disease, diabetes (categorised according to the most recent glycated haemoglobin [HbA1c] recorded in the 15 months prior to the first day of the pandemic wave), non-haematological and haematological cancer, chronic kidney disease or renal replacement therapy (categorised based on estimated glomerular filtration rates of ≥60 [absent], <60 and ≥45 [stage 3a],<45 and ≥30 [stage 3b], <30 and ≥15 [stage 4], and <15 [stage 5], respectively, or diagnostic codes indicative of dialysis and kidney transplant), chronic liver disease, stroke, dementia, other neurological disease (motor neurone disease, myasthenia gravis, multiple sclerosis, Parkinson’s disease, cerebral palsy, quadriplegia or hemiplegia, and progressive cerebellar disease), organ transplant (categorised as kidney transplant or other organ transplant), asplenia (splenectomy or a spleen dysfunction, including sickle cell disease), rheumatoid arthritis/lupus/psoriasis, learning disability, severe mental illness and other immunosuppressive conditions. STP of the patient’s general practice was included as an additional covariate to adjust for geographical variation in infection rates across the country.

#### Codelists and implementation

Information on all covariates were obtained from primary care records by searching TPP SystmOne records for specific coded data. Detailed information on compilation and sources for every individual codelist is available at https://codelists.opensafely.org/ and the lists are available for inspection and re-use by the broader research community.

#### Missing data

Missing data was expected in BMI, smoking and ethnicity. No missing data was expected in comorbidities as they were coded as present or absent. In the analysis, those with missing ethnicity were coded as unknown; those with missing BMI records were coded as non-obese; and those with missing smoking information were coded as non-smokers on the assumption that both obesity and smoking would be likely to be recorded if present. These assumptions have previously been tested in sensitivity analyses [1].

### Statistical methods

#### Absolute rate of death

Crude COVID-19-related death rates were calculated for the overall population and then stratified by subgroup (based on the covariates described above) for waves 1, 2 and 3. To account for age and sex differences in the various subgroups of the population, death rates were directly age- and sex-standardised to the European Standard Population using five-year age bands, except rates of death by age group (not standardised) and rates of death by sex (standardised for age). Confidence intervals were obtained taking the normal approximation to the binomial distribution [16]. Follow-up began on the first date of the wave and ended at the earliest occurrence of COVID-19-related death, death by other causes, or the end date of the wave. To capture changes in absolute COVID-19-related death rates over time, fold-changes in death rates (defined as the ratio between two death rates) were subsequently calculated for waves 2 and 3 compared to wave 1.

#### Relative hazard of death

For each of the clinical and demographic covariates of interest, a Cox proportional hazards model was fitted. Models were adjusted for age using restricted cubic splines with four knots, except for the estimation of relative hazard of death by age group; and adjusted for sex, except for estimation of relative hazard of death by sex; and stratified by STP region to account for regional differences in infection rates. Follow-up began on the first date of the wave and ended at the earliest occurrence of COVID-19-related death, death by other causes, or the end date of the wave. For clinical conditions, people without the clinical condition were used as the reference. The following reference values were used for the variables with more than two categories: age: 50-59 years, IMD: 5 (least deprived), smoking: never or unknown. All analyses were done separately for wave 1, wave 2 and wave 3. Proportional hazards assumptions were assessed by testing for a zero slope in the scaled Schoenfeld residuals and graphical methods. To capture changes in the relative hazard of COVID-19-related death over time, fold-changes in the relative hazard (defined as the ratio between relative hazards) were subsequently calculated for waves 2 and 3 compared to wave 1.

### Software and Reproducibility

Data management was performed using Python [3.8], with analysis carried out using R [4.0]. Code for data management and analysis, as well as codelists, are openly available online for inspection and re-use https://www.github.com/opensafely/covid_mortality_over_time.

## Results

### Study population

In total, 18.7, 18.8 and 18.7 million adults were included for waves 1, 2 and 3, respectively (Figure 1, Table 1, Figure A1-A2). In the final included study population, there were missing data for BMI (5,300,720 [28%]; 5,664,380 [30%]; 6,195,335 [33%]), smoking status (797,955 [4%]; 859,415 [5%]; 910,035 [5%]), and ethnicity (1,579,740 [8%]; 1,605,450 [9%]; 1,608,195 [9%]) for waves 1, 2 and 3, respectively. COVID-19-related death was recorded in linked death registration data for 15,570, 31,645, and 6,530 individuals in the study populations for waves 1, 2 and 3, respectively. Follow up time in the overall population was 3,474,600, 11,745,500 and 10,216,900 person-years for waves 1, 2 and 3 respectively. The percentage of people in the various population subgroups stayed approximately constant over time, except for some minor differences likely related to pandemic pressures. For example, the proportion of people with asthma not using oral steroids, increased from 2,713,510 in wave 1 (14.5%) to 2,758,900 in wave 2 (14.7%) and 2,837,475 in wave 3 (15.2%). A similar pattern was observed in people with diabetes without recent HbA1c measure, increasing from 205,375 in wave 1 (1.1%), to 302,775 in wave 2 (1.6%) and 389,665 in wave 3 (2.1%).

**Table 1.**
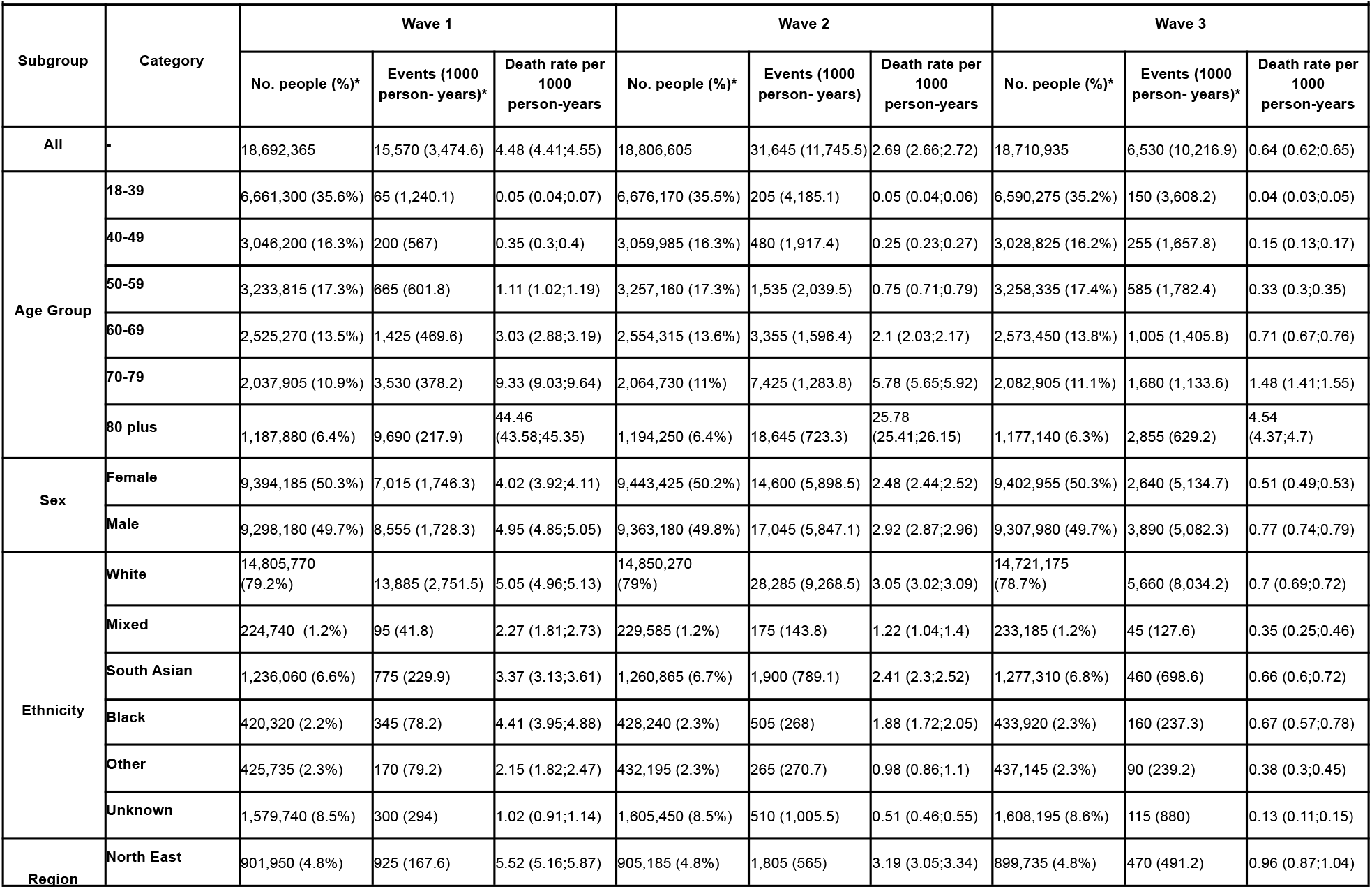
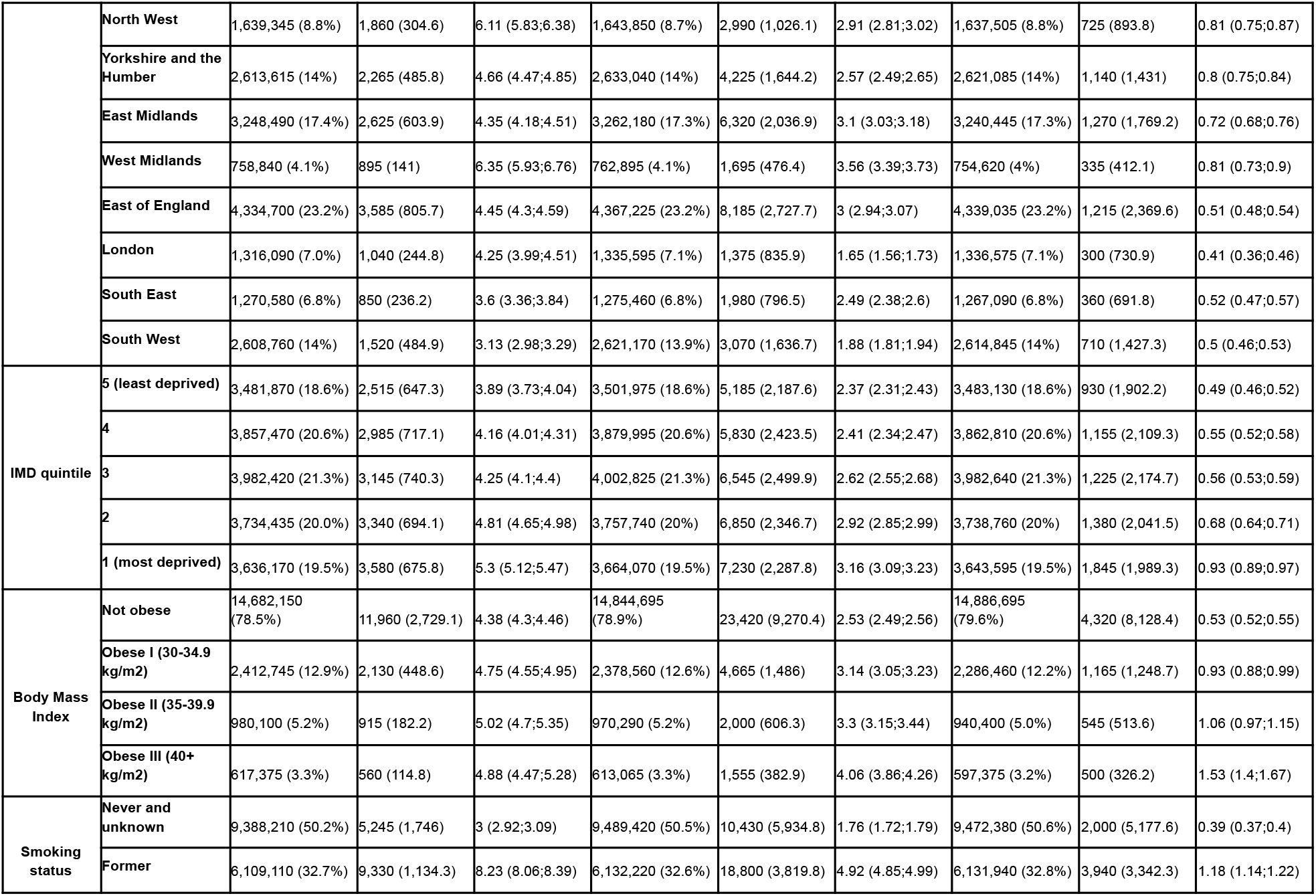
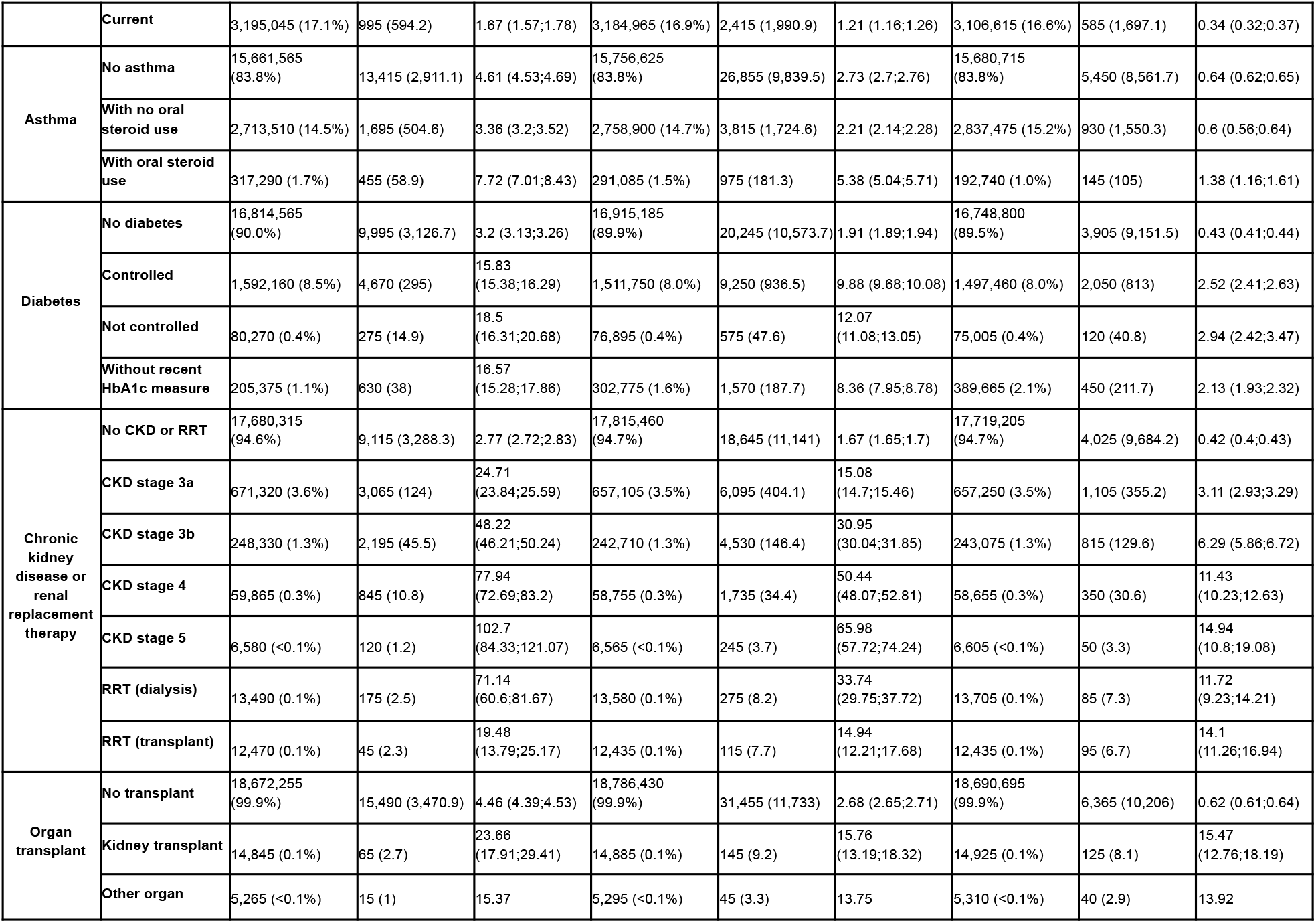
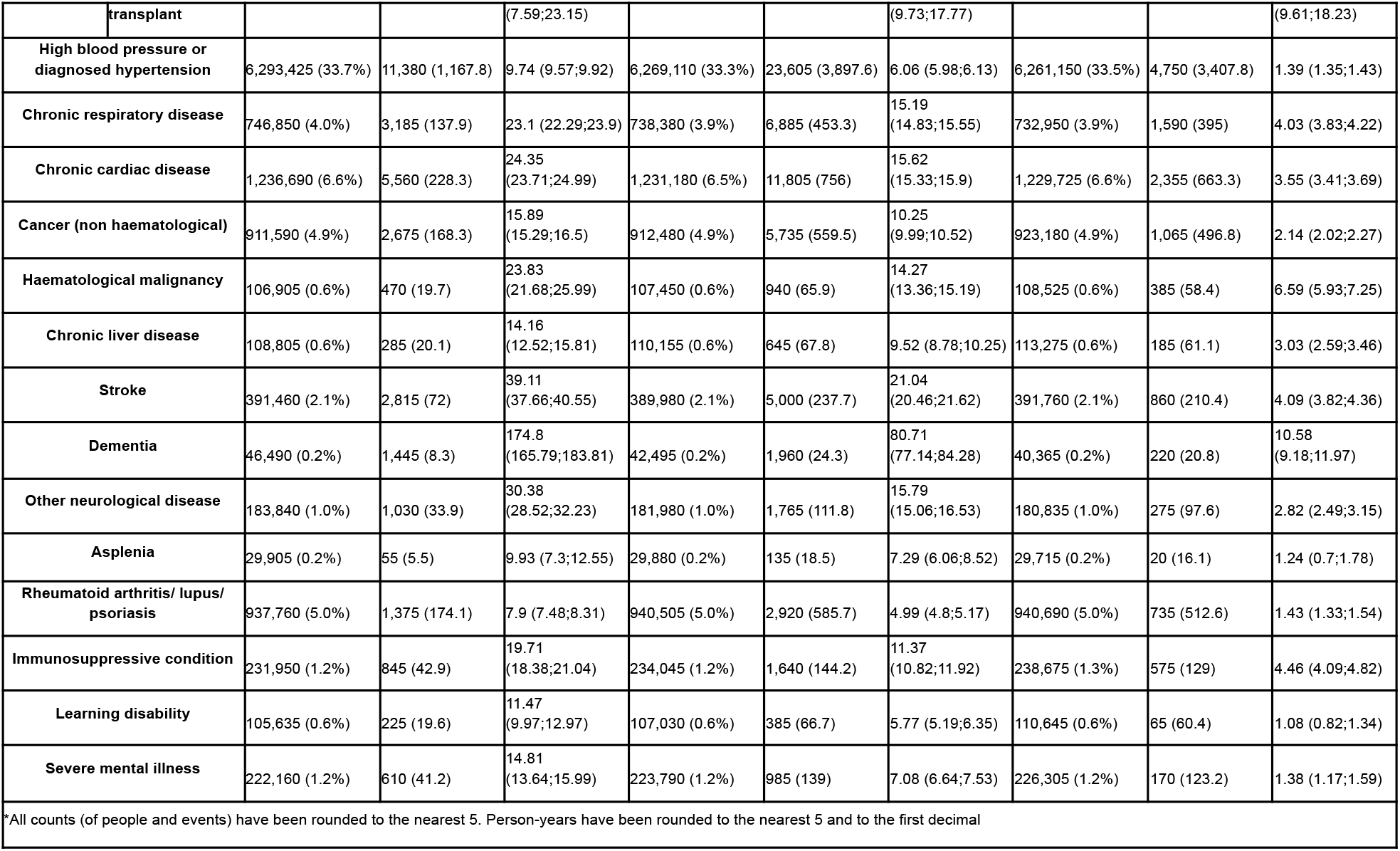
Number of people and COVID-19-related death rates in OpenSAFELY-TPP, stratified by demographic and clinical subgroups in the first three pandemic waves in England (wave 1: March 23 to May 30, 2020; wave 2: September 7, 2020 to April 24, 2021; and wave 3, delta: May 28 to December 14, 2021).

**Figure 1.**
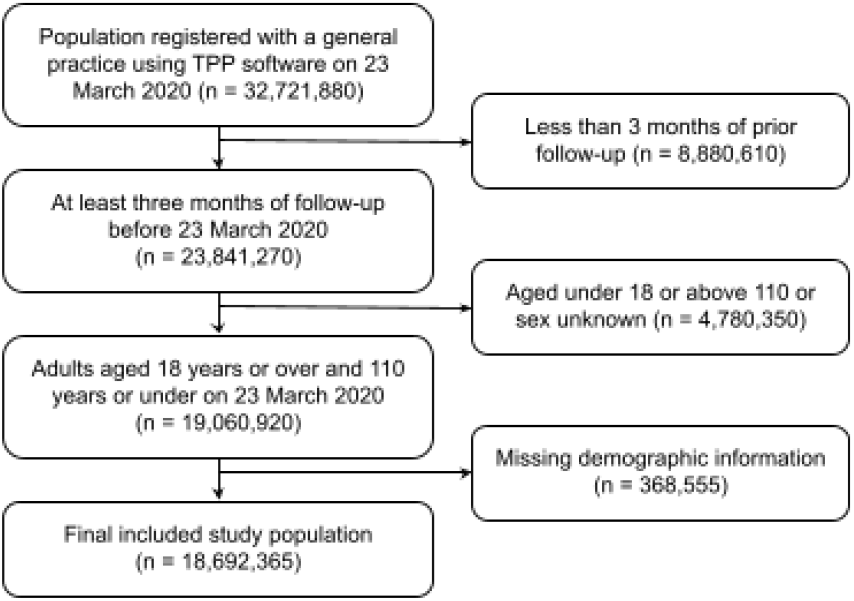
This diagram shows the number of people (*n*) excluded at different stages of cohort selection covering the first pandemic wave (23 March 2020 - 30 May 2020). All counts were rounded to the nearest five.

### Absolute rate of death

The crude COVID-19-related death rate in the overall population per 1,000 person-years decreased from 4.48 in wave 1 (95%CI 4.41;4.55) to 2.69 in wave 2 (95%CI 2.66;2.72) and 0.64 in wave 3 (95%CI 0.62;0.65). Crude death rates in all population subgroups were highest in wave 1, and decreased in the subsequent pandemic waves (Table 1).

The sex- and age-standardised COVID-19-related death rates were consistently lower in wave 2 and 3 compared to wave 1 (Figure 2, Table A1, Figure A3). The standardised death rates in the overall population per 1,000 person-years decreased from 4.58 in wave 1 (95%CI 4.50;4.65) to 2.77 in wave 2 (95%CI 2.74;2.80), and 0.65 in wave 3 (95%CI

**Figure 2.**
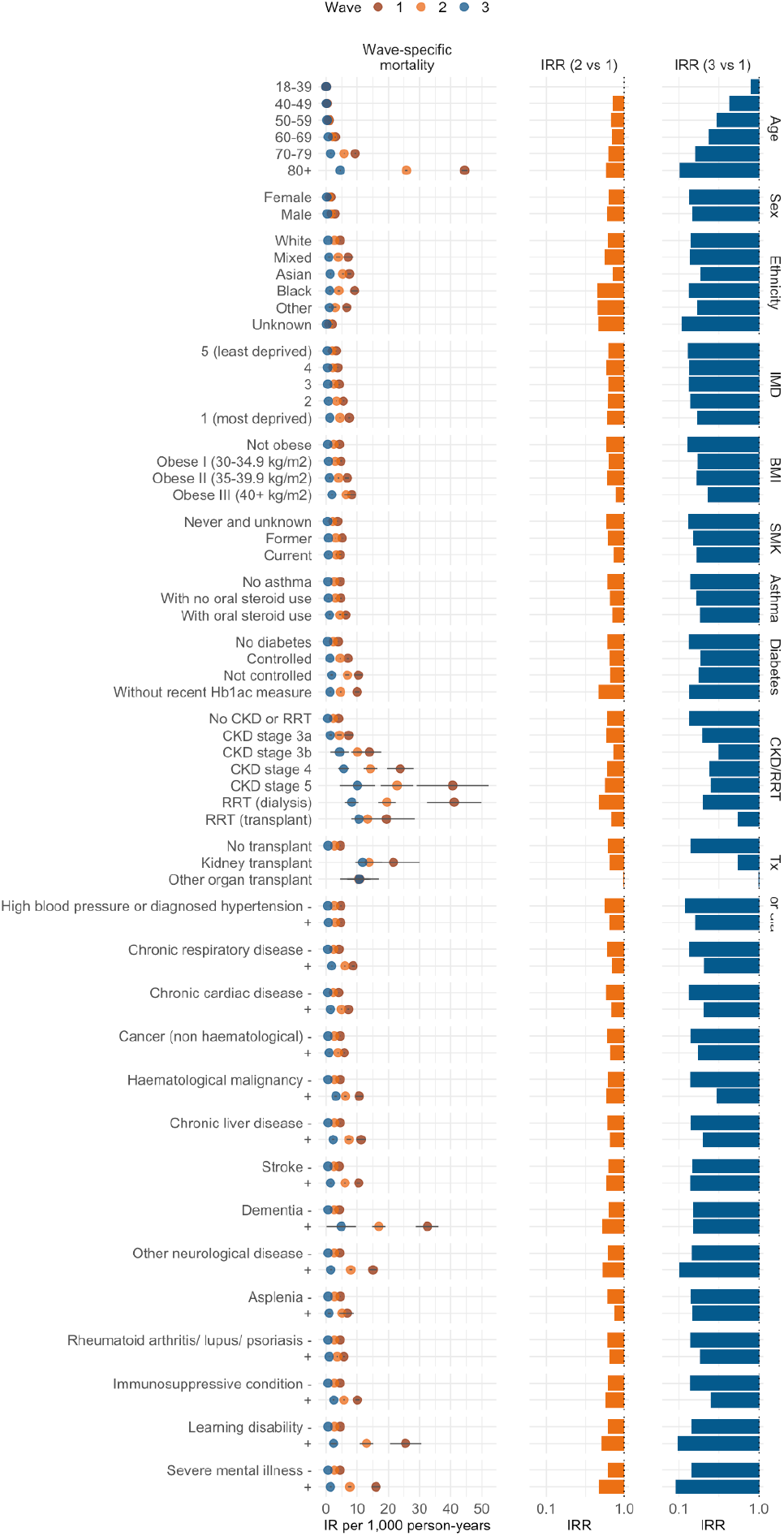
Sex- and age-standardised COVID-19-related death rates (IR) and 95% confidence intervals per 1,000 person-years in OpenSAFELY-TPP in the three pandemic waves (wave 1: March 23 to May 30, 2020; wave 2: September 7, 2020 to April 24, 2021; and wave 3, delta: May 28 to December 14, 2021). Models were standardised for age and sex using the European standard population except for the death rates by age group (not standardised) and death rates by sex (standardised by age). The two columns on the right present the fold-changes in death rate (IRR) of wave 2 vs 1 and wave 3 vs 1. Abbreviations: IMD, index of multiple deprivation; BMI, body mass index; SMK, smoking; CKD, chronic kidney disease; RRT, renal replacement therapy; Tx, transplant. The death rates presented in this figure can be found in table A1 of the Appendix.

0.63;0.67), representing an overall fold-change in death rate of 0.14 for wave 3 versus wave 1. Change in the death rates of wave 3 versus wave 1 was more pronounced in the older age groups compared to the younger age groups (Figure 2; fold-changes for wave 3 versus wave 1 ranging from 0.1 in the oldest age group to 0.8 in the youngest). In addition, a high fold-change for wave 3 versus wave 1 of 0.1 or lower was also seen for people with other neurological disease, severe mental illness, and learning disability (as compared to ≥ 0.13 for the other clinical and demographic subgroups). The decline in the standardised death rate across waves was attenuated in the organ transplant group. The standardised death rate per 1,000 person-years in the kidney transplant group was 21.59 in wave 1 (95%CI 13.15;30.04), 13.81 in wave 2 (95%CI 9.53;18.08) and 11.72 in wave 3 (95%CI 9.24;14.20), representing a fold-change of 0.54 for wave 3 versus wave 1. The standardised death rate per 1,000 person-years in the other organ transplant group was 10.75 in wave 1 (95%CI 4.45;17.05), 10.37 in wave 2 (95%CI 6.94;13.81) and 10.58 in wave 3 (95%CI 6.8;14.35), representing a fold-change of 0.98 for wave 3 versus wave 1. In addition, the change in death rates of wave 3 versus wave 1 was attenuated in people with severe obesity, kidney disease stage 3b-5, haematological malignancies and immunosuppressive conditions, represented by fold-changes of 0.23-0.54 for wave 3 versus wave 1 (as compared to ≤0.2 for the other clinical and demographic subgroups).

### Relative hazard of death

Patterns in subgroup-specific sex- and age-standardised relative hazards of death (stratified by STP region) were consistent throughout the three pandemic waves (Figure 3, Table A2). The relative hazard of death was higher in older people, male sex, ethnic minorities and in people with various medical conditions. Several clinical and demographic subgroups experienced a decline in the relative hazard of death (relative to the reference subgroup) over successive waves. The relative hazard of death for people aged 80 or over versus people aged 50-59 years old was 41.56 in wave 1 (95%CI 41.18;48.21), and 36.51 in wave 2 (95%CI 34.65;38.46), and decreased to 15.25 in wave 3 (95%CI 13.95;16.67), representing a fold-change of 0.37 for wave 3 versus wave 1. The relative hazard of death for people with versus without a learning disability was 8.73 in wave 1 (95%CI 7.65;9.96) and decreased to 6.97 in wave 2 (95%CI 6.30;7.71) and 3.95 in wave 3 (95%CI 3.09;5.05), representing a fold-change of 0.45 for wave 3 versus 1.

**Figure 3.**
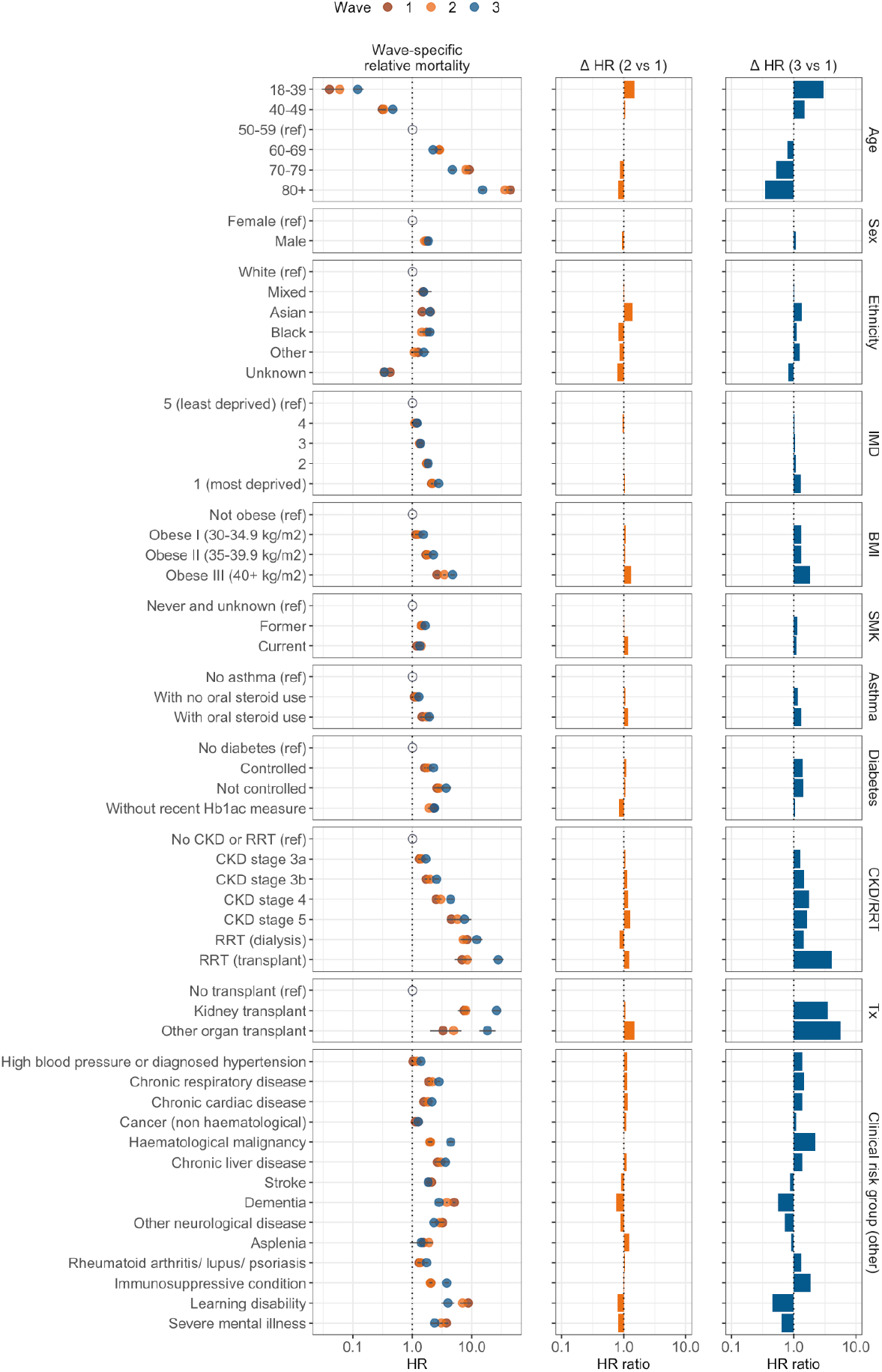
Relative hazard of COVID-19-related death (HR) and 95% confidence intervals in OpenSAFELY-TPP in the three pandemic waves (wave 1: March 23 to May 30, 2020; wave 2: September 7, 2020 to April 24, 2021; and wave 3, delta: May 28 to December 14, 2021). Models were adjusted for age using a 4-knot cubic spline, except for estimation of age group relative hazard of death; and adjusted for sex, except for estimation of sex group relative hazard of death; and stratified by region. The two columns on the right present the ratio of the relative hazard of death (fold-change: Δ HR) of wave 2 vs 1 and wave 3 vs 1. Abbreviations: IMD, index of multiple deprivation; BMI, body mass index; SMK, smoking; CKD, chronic kidney disease; RRT, renal replacement therapy; Tx, transplant. The relative hazards of death presented in this figure can be found in table A2 of the Appendix.

In some demographic subgroups, the relative hazard of death stayed comparably constant across successive waves (compared to the decrease in relative hazard of death in e.g. people of older age). The relative hazard of death for male versus female sex was 1.72 in wave 1 (95%CI 1.66;1.77), decreased to 1.61 in wave 2 (95%CI 1.57;1.64), and increased to 1.86 in wave 3 (95%CI 1.77;1.96), representing a fold-change of 1.08 for wave 3 versus wave 1. A similar pattern was seen in the relative hazard of death for ethnicity and IMD. However, an exception included the relative hazard of death for Asian versus White ethnicity, increasing from 1.47 in wave 1 (95%CI 1.36;1.59) to 2.02 in wave 2 (95%CI 1.92;2.12), and 1.99 in wave 3 (95%CI 1.80;2.20), representing a fold-change of 1.35 for wave 3 versus wave 1. Another exception was the relative hazard of death for IMD 1 (most deprived) versus IMD 5 (least deprived), going from 2.21 in wave 1 (95%CI 2.01;2.24) to 2.19 in wave 2 (95%CI 2.11;2.27), and 2.77 in wave 3 (95%CI 2.56;3.01), representing a fold-change of 1.31 for wave 3 versus wave 1.

What is more, the relative hazard of death was higher in wave 3 compared to wave 1 in several population subgroups, most notably in people with kidney disease and organ transplants recipients. The relative hazard of death for CKD stage 5 versus no kidney disease increased from 4.52 in wave 1 (95%CI 3.77;5.42) to 7.47 in wave 3 (95%CI 5.61;9.95), representing a fold-change of 1.65, while the relative hazard of death for people on dialysis versus no kidney disease increased from 8.30 in wave 1 (95%CI 7.15;9.63) to 12.09 in wave 3 (95%CI 9.77;14.96), representing a fold-change of 1.55. The relative hazard of death for kidney transplant versus no transplant increased from 7.37 in wave 1 (95%CI 5.78;9.41) to 26.33 in wave 3 (95%CI 22.00;31.50), representing a fold-change of 3.57, while the relative hazard of death for other organ transplant versus no transplant increased from 3.27 (95%CI 1.97;5.43) in wave 1 to 18.34 in wave 3 (95%CI 13.39;25.14), representing a fold-change of 5.61. Other population subgroups with high fold-changes of relative hazard of death in wave 3 included 18-39 year olds (likely reflecting delayed eligibility for or low uptake of vaccination) and individuals with severe obesity, haematological malignancy, and immunosuppressive conditions (Figure 3; fold-changes of 1.8-3 for wave 3 vs wave 1)

## Discussion

### Summary

In this observational study in 18 million adults in England, we found that overall COVID-19-related death rates decreased from waves 1 to 2 to 3. A high decrease in death rates was seen in groups of older age and people with neurological disease, learning disability or severe mental illness. Conversely, decreases in COVID-19 death rates across successive waves were substantially attenuated in several groups: organ transplant recipients; people with CKD including those receiving dialysis treatment; people with haematological malignancies; and those with conditions associated with immunosuppression. Consequently, relative hazards of COVID-19-related death in individuals with versus without these clinical conditions rose by 2–6 fold in wave 3 compared to wave 1. These groups are more likely to experience impaired vaccine effectiveness [17], [18].

The observed decrease in COVID-19-related death rates is likely to be driven in part by the vaccination programme introduced on 8 December 2020 midway through wave 2, with the oldest and most vulnerable groups prioritised first. Absolute rate of mortality fell by 90% in wave 3 compared to wave 1 among the over 80s who were eligible for vaccination at the outset of the vaccination programme, whereas the corresponding decline was 20% in those aged 18-39 where only the most vulnerable people were eligible for vaccination at the start of the programme [19], [20]. Other mechanisms likely to have driven the decrease in COVID-19-related death rates are the improved clinical management of COVID-19 disease over time [21], [22] and coronavirus lockdowns and various restrictions [23], [24].

When considering relative hazards of COVID-19-related death across subgroups, in age- and sex-adjusted models, higher hazards of death (relative hazards of death >1 in each pandemic wave) were observed for individuals with clinical conditions compared to those without the condition. For conditions such as kidney disease, greater severity of disease was associated with higher relative hazards of death. As expected, the relative hazard of death was higher with increasing age, although relative hazards for 80 year olds versus 50-59 year olds decreased by 66% from waves 1 to 3 (relative hazard of death of 45 and 15, respectively). Consistent patterns of higher relative hazards of death were observed in association with higher social deprivation, male sex and non-White ethnicity across successive waves, with mostly small fold-changes in relative hazard of death accross succesive waves (1.0–1.4-fold rise in relative hazard of death of wave 3 versus wave 1). Several clinical subgroups had an increased relative hazard of death across successive waves, including organ transplant recipients (3.6–5.6-fold rise in relative hazard of death of wave 3 versus wave 1), CKD and dialysis patients (1.3–1.7-fold rise), patients with haematological malignancy (2.2-fold rise), individuals with severe obesity (1.8-fold rise), and patients with immunosuppressive conditions (1.8-fold rise). These increases in the relative hazards are in keeping with the attenuated declines in absolute death rates seen in wave 3 in these clinical subgroups.

### Strengths and weaknesses

This study used large-scale, routinely-collected primary care records, linked to death registry data. This allowed us to describe a substantial proportion of the English population in the first three pandemic waves and to describe changes over time in population-level COVID-19-related mortality in population subgroups based on clinical and demographic characteristics.

We acknowledge several important limitations to these results. Classification error in ICD codes used to identify COVID-19-related deaths might have occured. COVID-19-related death may have been misclassified as being due to other causes, particularly early in the first pandemic wave where mass testing was not available. In addition, ascertainment of certain clinical conditions may have been imperfect, as it relies on appropriate and timely clinical coding in the primary care record. A patient must present to the GP, or have information fed back properly from secondary care settings, to be coded correctly. This issue may be more pronounced for less severe illnesses or early-stage conditions. Small declines were observed in the percentage of people with asthma and diabetes over the period of the pandemic, which may reflect a reduction in health care utilisation as a consequence of the COVID-19 pandemic, rather than a real change in disease incidence.

This study cannot disentangle the pathway from infection to disease to death since the ascertainment of infection status is not consistently reliable in routinely-collected health data and changes with underlying community infection rates, test availability and access (particularly early in the pandemic), health-seeking behaviour and symptoms. We therefore did not quantify the risk of COVID-19 death given SARS-CoV-2 infection or the risk of infection separately, and reported death rates should not be interpreted in this way. The risk of infection is affected by contact patterns of individuals and their local infection incidence. Public health guidance and support for contact-reducing interventions (“shielding”) as well as public perception of risk was changing across the study period, and therefore the risk of infection for clinically vulnerable individuals may have varied from wave to wave. Additionally, the risk of infection is affected by SARS-CoV-2 variants, with Delta (wave 3) being associated with increased transmissibility compared to the Alpha variant (wave 2) [25].

Despite our sample of patients registered at practices using TPP software under-representing some geographic areas, particularly London and the North West, absolute rates presented in this study should be considered to represent rates across the whole of England [26].

### Findings in Context

Previous studies of COVID-19-related mortality in demographic and clinical subgroups have shown differences in risks consistent with those presented here, including an increased risk in groups of older age, male sex, social deprivation, non-White ethnicity, and clinical conditions such as kidney disease, organ transplant, and learning disability [1], [15]. Our findings highlight the extent to which death rates have changed over the course of the pandemic. Notably, during wave 3, when primary vaccination using ChAdOx1-S or BNT162b2 had been offered to high-risk groups in England, death rates in groups of older age, learning disability, and severe mental illness were markedly attenuated compared to waves 1 and 2. This decline is in line with the high effectiveness of primary vaccination against COVID-19-related death [27]. It is also possible that the Delta variant (dominant in wave 3) had lower consequent mortality while being more infectious compared to Alpha (dominant in wave 2) [25] and more likely to escape vaccination [28].

An impaired response to primary COVID-19 vaccination has been reported in immunocompromised populations, including organ transplant recipients, dialysis patients, haematologic cancer patients, and individuals receiving immunosuppressive therapy [17], [18], [29]. Notably, these studies have typically focused on post-vaccination antibody levels or effectiveness against symptomatic or severe disease. In the present study, we highlight the excess COVID-19-related mortality that persists among clinical subgroups that have been linked with impaired primary vaccine response. Compared with the overall population, these groups experienced more modest declines in absolute death rates in wave 3 (after widespread vaccine implementation) compared to waves 1 and 2.

### Policy Implications and Interpretation

Some of the demographic inequalities in COVID-19 mortality burden observed here are mirrored by inequality in COVID-19 vaccine coverage [20], most notably ethnicity. Given the demonstrated efficacy of COVID vaccines in reducing COVID-19-related death [27], it seems likely that better targeting of vaccines to these groups would have helped to reduce their mortality burden in second and third waves.

The same benefit may be more challenging to obtain for patient groups where vaccine efficacy may be lower, such as conditions associated with lower immune response. Despite such groups being targeted for early vaccination in many cases, absolute risks have persisted between waves. Our findings provide an evidence-base to inform UK public health policy for protecting these vulnerable patients.

## Conclusions

Despite population-level reductions in COVID-19-related mortality, there are still persistent inequalities among different clinical and socio-economic groups. Further, certain clinical subgroups remain highly vulnerable, particularly people with conditions associated with impaired immune response.

## Data Availability

Access to the underlying identifiable and potentially re-identifiable pseudonymised electronic health record data is tightly governed by various legislative and regulatory frameworks, and restricted by best practice.

## Abbreviations

CI: confidence interval
SARS-CoV-2: severe acute respiratory syndrome coronavirus 2
COVID-19: Coronavirus disease 2019
BMI: body mass index
CKD: chronic kidney disease
RRT: renal replacement therapy
IMD: index of multiple deprivation
STP: Sustainability and Transformation Partnership region

## Acknowledgements

We are very grateful for all the support received from the TPP Technical Operations team throughout this work, and for generous assistance from the information governance and database teams at NHS England / NHSX.

## Information governance and ethical approval

NHS England is the data controller for OpenSAFELY-TPP; TPP is the data processor; all study authors using OpenSAFELY have the approval of NHS England. This implementation of OpenSAFELY is hosted within the TPP environment which is accredited to the ISO 27001 information security standard and is NHS IG Toolkit compliant.[30]

Patient data has been pseudonymised for analysis and linkage using industry standard cryptographic hashing techniques; all pseudonymised datasets transmitted for linkage onto OpenSAFELY are encrypted; access to the platform is via a virtual private network (VPN) connection, restricted to a small group of researchers; the researchers hold contracts with NHS England and only access the platform to initiate database queries and statistical models; all database activity is logged; only aggregate statistical outputs leave the platform environment following best practice for anonymisation of results such as statistical disclosure control for low cell counts.[31]

The OpenSAFELY research platform adheres to the obligations of the UK General Data Protection Regulation (GDPR) and the Data Protection Act 2018. In March 2020, the Secretary of State for Health and Social Care used powers under the UK Health Service (Control of Patient Information) Regulations 2002 (COPI) to require organisations to process confidential patient information for the purposes of protecting public health, providing healthcare services to the public and monitoring and managing the COVID-19 outbreak and incidents of exposure; this sets aside the requirement for patient consent.[32] This was extended in July 2022 for the NHS England OpenSAFELY COVID-19 research platform.[33] In some cases of data sharing, the common law duty of confidence is met using, for example, patient consent or support from the Health Research Authority Confidentiality Advisory Group.[34]

Taken together, these provide the legal bases to link patient datasets on the OpenSAFELY platform. GP practices, from which the primary care data are obtained, are required to share relevant health information to support the public health response to the pandemic, and have been informed of the OpenSAFELY analytics platform.

This study was approved by the Health Research Authority (REC reference 20/LO/0651) and by the LSHTM Ethics Board (reference 21863).

## Transparency

The guarantor affirms that the manuscript is an honest, accurate, and transparent account of the study being reported; that no important aspects of the study have been omitted; and that any discrepancies from the study as planned have been explained.

## Data access and verification

Access to the underlying identifiable and potentially re-identifiable pseudonymised electronic health record data is tightly governed by various legislative and regulatory frameworks, and restricted by best practice. The data in OpenSAFELY is drawn from General Practice data across England where TPP is the Data Processor. TPP developers (CB, JC, JP, FH, and SH) initiate an automated process to create pseudonymised records in the core OpenSAFELY database, which are copies of key structured data tables in the identifiable records. These are linked onto key external data resources that have also been pseudonymised via SHA-512 one-way hashing of NHS numbers using a shared salt. Bennett Institute for Applied Data Science developers and PIs (BG, CEM, SCJB, AJW, WJH, DE, PI, SD, GH, BBC, RMS, ID, TW, TO, SM, CLS, LB and EJW) holding contracts with NHS England have access to the OpenSAFELY pseudonymised data tables as needed to develop the OpenSAFELY tools. These tools in turn enable researchers with OpenSAFELY Data Access Agreements to write and execute code for data management and data analysis without direct access to the underlying raw pseudonymised patient data, and to review the outputs of this code. All code for the full data management pipeline—from raw data to completed results for this analysis—and for the OpenSAFELY platform as a whole is available for review at github.com/OpenSAFELY.

The data management and analysis code for this paper was led by LN and contributed to by CDA, LF, WJH and AJW.

## Appendix

**Figure A1.**
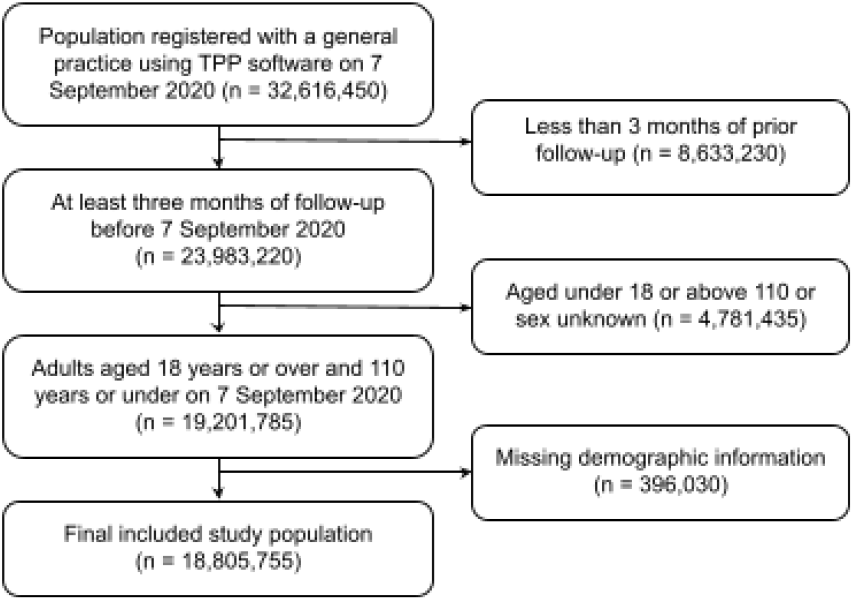
This diagram shows the number of people (*n*) excluded at different stages of cohort selection covering the second pandemic wave (7 September 2020 - 24 April 2021). All counts were rounded to the nearest five.

**Figure A2.**
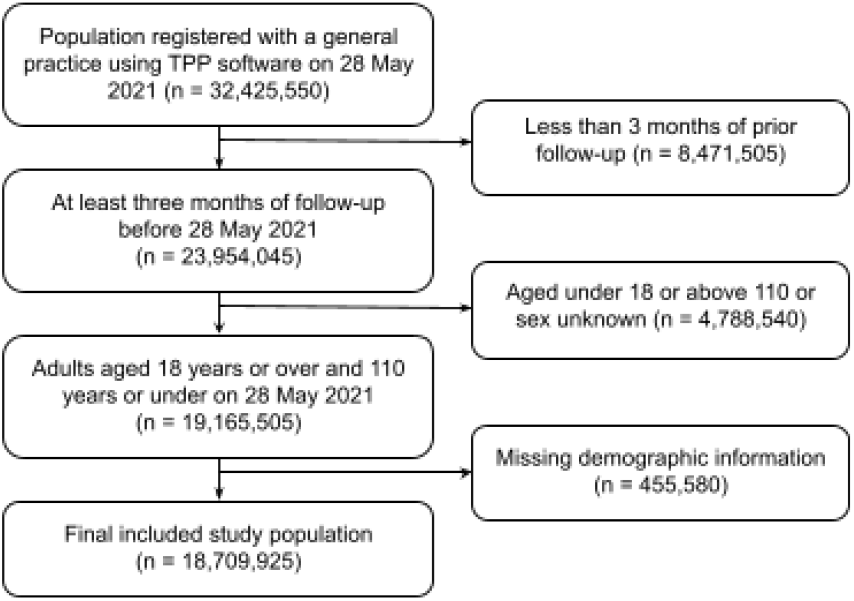
This diagram shows the number of people (*n*) excluded at different stages of cohort selection covering the third pandemic wave (28 May 2021 - 14 December 2021). All counts were rounded to the nearest five.

**Figure A3.**
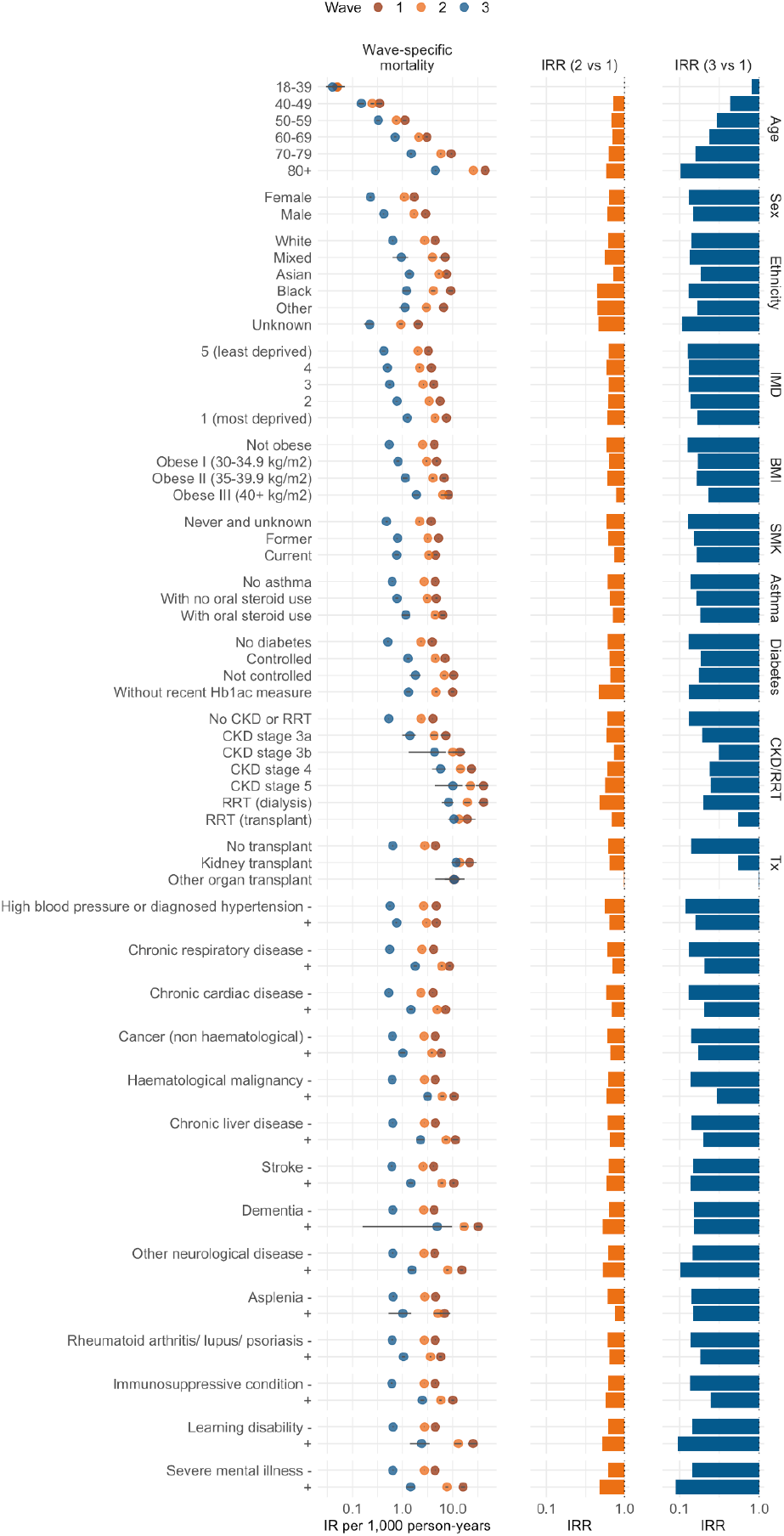
Sex- and age-standardised COVID-19-related death rates and 95% confidence intervals per 1,000 person-years in OpenSAFELY-TPP in the three pandemic waves (wave 1: March 23 to May 30, 2020; wave 2: September 7, 2020 to April 24, 2021; and wave 3, delta: May 28 to December 14, 2021). Models were standardised for age and sex using the European standard population except for the death rates by age group (not standardised) and death rates by sex (standardised by age). The two columns on the right present the fold-change in death rate of wave 2 vs 1 and wave 3 vs 1. Abbreviations: IMD, index of multiple deprivation; BMI, body mass index; SMK, smoking; CKD, chronic kidney disease; RRT, renal replacement therapy; Tx, transplant. The death rates presented in this figure can be found in table A1 of the Appendix.

**Table A1.** Sex- and age-standardised COVID-19-related death rates and 95% confidence intervals per 1,000 person-years in OpenSAFELY-TPP in the three pandemic waves (wave 1: March 23 to May 30, 2020; wave 2: September 7, 2020 to April 24, 2021; and wave 3, delta: May 28 to December 14, 2021).

| Subgroup | Level | COVID-19-related death rate* per 1000 person-years (95% CI) |  |  |
| --- | --- | --- | --- | --- |
|  |  | Wave 1 | Wave 2 | Wave 3 |
| All | - | 4.58 (4.50;4.65) | 2.77 (2.74;2.80) | 0.65 (0.63;0.67) |
| Age Group | 18-39 | 0.05 (0.04;0.07) | 0.05 (0.04;0.06) | 0.04 (0.03;0.05) |
|  | 40-49 | 0.35 (0.3;0.4) | 0.25 (0.23;0.27) | 0.15 (0.13;0.17) |
|  | 50-59 | 1.11 (1.02;1.19) | 0.75 (0.71;0.79) | 0.33 (0.30;0.35) |
|  | 60-69 | 3.03 (2.88;3.19) | 2.1 (2.03;2.17) | 0.71 (0.67;0.76) |
|  | 70-79 | 9.33 (9.03;9.64) | 5.78 (5.65;5.92) | 1.48 (1.41;1.55) |
|  | 80 plus | 44.46 (43.58;45.35) | 25.78 (25.41;26.15) | 4.54 (4.37;4.7) |
| Sex | Female | 1.72 (1.68;1.76) | 1.08 (1.06;1.1) | 0.23 (0.22;0.24) |
|  | Male | 2.86 (2.8;2.92) | 1.69 (1.66;1.72) | 0.42 (0.41;0.43) |
| Ethnicity | White | 4.5 (4.43;4.58) | 2.75 (2.71;2.78) | 0.64 (0.62;0.65) |
|  | Mixed | 7.06 (5.42;8.69) | 3.92 (3.26;4.58) | 0.95 (0.63;1.28) |
|  | Asian | 7.48 (6.91;8.05) | 5.33 (5.07;5.59) | 1.37 (1.23;1.51) |
|  | Black | 9.18 (8.10;10.27) | 4.09 (3.69;4.48) | 1.21 (1.00;1.43) |
|  | Other | 6.61 (5.53;7.7) | 2.96 (2.57;3.35) | 1.11 (0.85;1.37) |
|  | Unknown | 2.03 (1.79;2.28) | 0.93 (0.85;1.02) | 0.22 (0.17;0.26) |
| Region | North East | 5.66 (5.28;6.04) | 3.43 (3.26;3.59) | 0.99 (0.90;1.08) |
|  | North West | 6.05 (5.77;6.34) | 2.91 (2.80;3.02) | 0.79 (0.73;0.85) |
|  | Yorkshire and the Humber | 5.12 (4.90;5.34) | 2.83 (2.74;2.92) | 0.85 (0.80;0.90) |
|  | East Midlands | 4.47 (4.29;4.65) | 3.2 (3.12;3.28) | 0.73 (0.69;0.77) |
|  | West Midlands | 7.25 (6.77;7.74) | 4.1 (3.9;4.31) | 0.91 (0.81;1.01) |
|  | East of England | 4.49 (4.34;4.64) | 3.01 (2.94;3.08) | 0.52 (0.49;0.55) |
|  | London | 8.12 (7.6;8.63) | 3.07 (2.9;3.24) | 0.75 (0.66;0.84) |
|  | South East | 3.21 (2.98;3.43) | 2.23 (2.12;2.33) | 0.48 (0.43;0.54) |
|  | South West | 2.59 (2.46;2.73) | 1.57 (1.51;1.63) | 0.42 (0.39;0.45) |
| IMD quintile | 5 (least deprived) | 3.28 (3.15;3.41) | 2.02 (1.96;2.07) | 0.42 (0.39;0.45) |
|  | 4 | 3.74 (3.6;3.88) | 2.18 (2.12;2.24) | 0.5 (0.47;0.53) |
|  | 3 | 4.17 (4.02;4.32) | 2.57 (2.51;2.63) | 0.55 (0.52;0.58) |
|  | 2 | 5.56 (5.37;5.76) | 3.41 (3.32;3.49) | 0.77 (0.73;0.81) |
|  | 1 (most deprived) | 7.45 (7.19;7.70) | 4.44 (4.34;4.55) | 1.25 (1.19;1.31) |
| Body Mass Index | Not obese | 4.29 (4.21;4.37) | 2.52 (2.49;2.55) | 0.54 (0.53;0.56) |
|  | Obese I (30-34.9 kg/m2) | 4.79 (4.56;5.02) | 3.01 (2.91;3.11) | 0.81 (0.76;0.86) |
|  | Obese II (35-39.9 kg/m2) | 6.77 (6.14;7.40) | 4.01 (3.78;4.24) | 1.13 (1.01;1.24) |
|  | Obese III (40+ kg/m2) | 8.24 (7.02;9.46) | 6.4 (5.8;7.01) | 1.89 (1.62;2.16) |
| Smoking status | Never and unknown | 3.71 (3.59;3.82) | 2.17 (2.12;2.22) | 0.48 (0.46;0.5) |
|  | Former | 5.21 (5.1;5.32) | 3.18 (3.13;3.22) | 0.79 (0.76;0.81) |
|  | Current | 4.57 (4.21;4.94) | 3.34 (3.16;3.52) | 0.76 (0.68;0.85) |
| Asthma | No asthma | 4.5 (4.42;4.58) | 2.69 (2.66;2.72) | 0.62 (0.61;0.64) |
|  | With no oral steroid use | 4.73 (4.49;4.97) | 3.08 (2.97;3.18) | 0.77 (0.72;0.82) |
|  | With oral steroid use | 6.35 (5.73;6.98) | 4.47 (4.17;4.77) | 1.16 (0.96;1.36) |
| Diabetes** | No diabetes | 3.88 (3.8;3.96) | 2.34 (2.3;2.37) | 0.51 (0.49;0.52) |
|  | Controlled | 7.04 (6.79;7.28) | 4.51 (4.38;4.63) | 1.29 (1.21;1.37) |
|  | Not controlled | 10.36 (9.05;11.68) | 6.83 (6.19;7.47) | 1.82 (1.36;2.28) |
|  | Without recent HbA1c measure | 9.96 (9.15;10.77) | 4.66 (4.41;4.90) | 1.32 (1.18;1.46) |
| Chronic kidney disease or renal replacement therapy | No CKD or RRT | 4 (3.91;4.09) | 2.36 (2.32;2.39) | 0.53 (0.51;0.54) |
|  | CKD stage 3a | 7.25 (5.75;8.75) | 4.27 (3.56;4.98) | 1.4 (0.99;1.81) |
|  | CKD stage 3b | 13.88 (9.98;17.78) | 10.04 (8.07;12.01) | 4.34 (1.3;7.38) |
|  | CKD stage 4 | 23.83 (19.53;28.14) | 14.21 (12.04;16.38) | 5.67 (3.91;7.44) |
|  | CKD stage 5 | 40.63 (29.12;52.14) | 22.77 (17.62;27.91) | 10.03 (4.37;15.68) |
|  | RRT (dialysis) | 41.11 (32.47;49.76) | 19.53 (16.69;22.38) | 8.25 (6.06;10.44) |
|  | RRT (transplant) | 19.39 (10.22;28.55) | 13.32 (8.95;17.68) | 10.52 (8.1;12.95) |
| Organ transplant | No transplant | 4.56 (4.48;4.63) | 2.76 (2.72;2.79) | 0.64 (0.62;0.65) |
|  | Kidney transplant | 21.59 (13.15;30.04) | 13.81 (9.53;18.08) | 11.72 (9.24;14.2) |
|  | Other organ transplant | 10.75 (4.45;17.05) | 10.37 (6.94;13.81) | 10.58 (6.8;14.35) |
| High blood pressure or diagnosed hypertension | No | 4.74 (4.59;4.89) | 2.63 (2.57;2.69) | 0.56 (0.53;0.59) |
|  | Yes | 4.74 (4.64;4.83) | 2.99 (2.95;3.04) | 0.76 (0.73;0.78) |
| **Chronic respiratory disease** | **No** | 4.13 (4.06;4.21) | 2.45 (2.42;2.48) | 0.55 (0.53;0.56) |
|  | **Yes** | 8.67 (8.22;9.12) | 6.05 (5.81;6.29) | 1.8 (1.65;1.95) |
| **Chronic cardiac disease** | **No** | 4.03 (3.94;4.11) | 2.33 (2.3;2.36) | 0.53 (0.51;0.55) |
|  | **Yes** | 7.18 (6.89;7.46) | 4.91 (4.76;5.06) | 1.46 (1.33;1.6) |
| **Cancer (non haematological)** | **No** | 4.51 (4.43;4.59) | 2.68 (2.64;2.71) | 0.63 (0.62;0.65) |
|  | **Yes** | 5.87 (5.54;6.20) | 3.85 (3.66;4.04) | 1.01 (0.88;1.14) |
| **Haematological malignancy** | **No** | 4.51 (4.44;4.59) | 2.73 (2.7;2.76) | 0.62 (0.61;0.64) |
|  | **Yes** | 10.59 (9.41;11.76) | 6.2 (5.71;6.69) | 3.15 (2.75;3.56) |
| **Chronic liver disease** | **No** | 4.53 (4.45;4.60) | 2.73 (2.70;2.77) | 0.64 (0.62;0.65) |
|  | **Yes** | 11.25 (9.75;12.74) | 7.34 (6.70;7.99) | 2.28 (1.91;2.66) |
| **Stroke** | **No** | 4.15 (4.08;4.23) | 2.57 (2.53;2.60) | 0.61 (0.59;0.63) |
|  | **Yes** | 10.43 (9.88;10.98) | 6.07 (5.70;6.43) | 1.45 (1.28;1.63) |
| **Dementia** | **No** | 4.24 (4.17;4.31) | 2.64 (2.61;2.67) | 0.64 (0.62;0.65) |
|  | **Yes** | 32.50 (28.84;36.16) | 16.94 (14.80;19.07) | 4.91 (0.16;9.66) |
| **Other neurological disease** | **No** | 4.37 (4.30;4.44) | 2.67 (2.64;2.70) | 0.64 (0.62;0.65) |
|  | **Yes** | 15.06 (14.04;16.07) | 7.92 (7.51;8.32) | 1.53 (1.33;1.74) |
| **Asplenia** | **No** | 4.57 (4.50;4.65) | 2.76 (2.73;2.80) | 0.65 (0.63;0.67) |
|  | **Yes** | 6.84 (4.82;8.87) | 5.09 (4.18;6.00) | 1.01 (0.52;1.50) |
| **Rheumatoid arthritis/ lupus/ psoriasis** | **No** | 4.49 (4.41;4.56) | 2.71 (2.67;2.74) | 0.62 (0.60;0.64) |
|  | **Yes** | 5.72 (5.40;6.04) | 3.61 (3.47;3.74) | 1.04 (0.96;1.12) |
| **Immunosuppressive condition** | **No** | 4.46 (4.38;4.53) | 2.7 (2.67;2.73) | 0.61 (0.59;0.63) |
|  | **Yes** | 10.02 (9.28;10.77) | 5.75 (5.45;6.05) | 2.49 (2.25;2.72) |
| **Learning disability** | **No** | 4.52 (4.45;4.59) | 2.74 (2.71;2.77) | 0.65 (0.63;0.66) |
|  | **Yes** | 25.46 (20.40;30.53) | 13 (10.83;15.18) | 2.41 (1.41;3.41) |
| **Severe mental illness** | **No** | 4.45 (4.38;4.52) | 2.72 (2.68;2.75) | 0.64 (0.63;0.66) |
|  | **Yes** | 16.05 (14.64;17.47) | 7.63 (7.10;8.16) | 1.44 (1.19;1.69) |

\* Models were standardised for age and sex using the European standard population except for the death rates by age group (not standardised) and death rates by sex (standardised by age).
\*\* HbA1c classification based on the latest measure within 15 months before baseline.

**Table A2.**
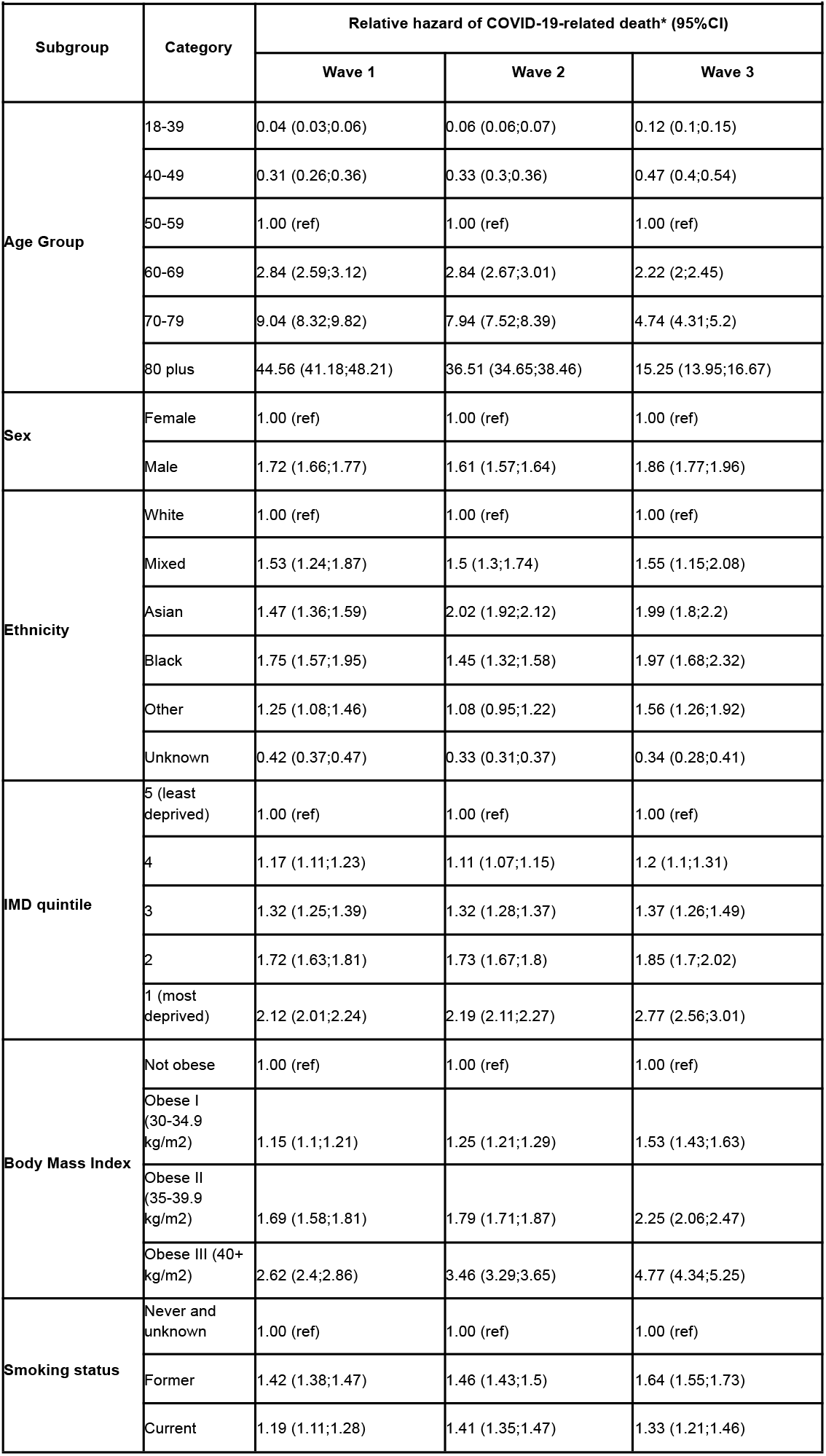
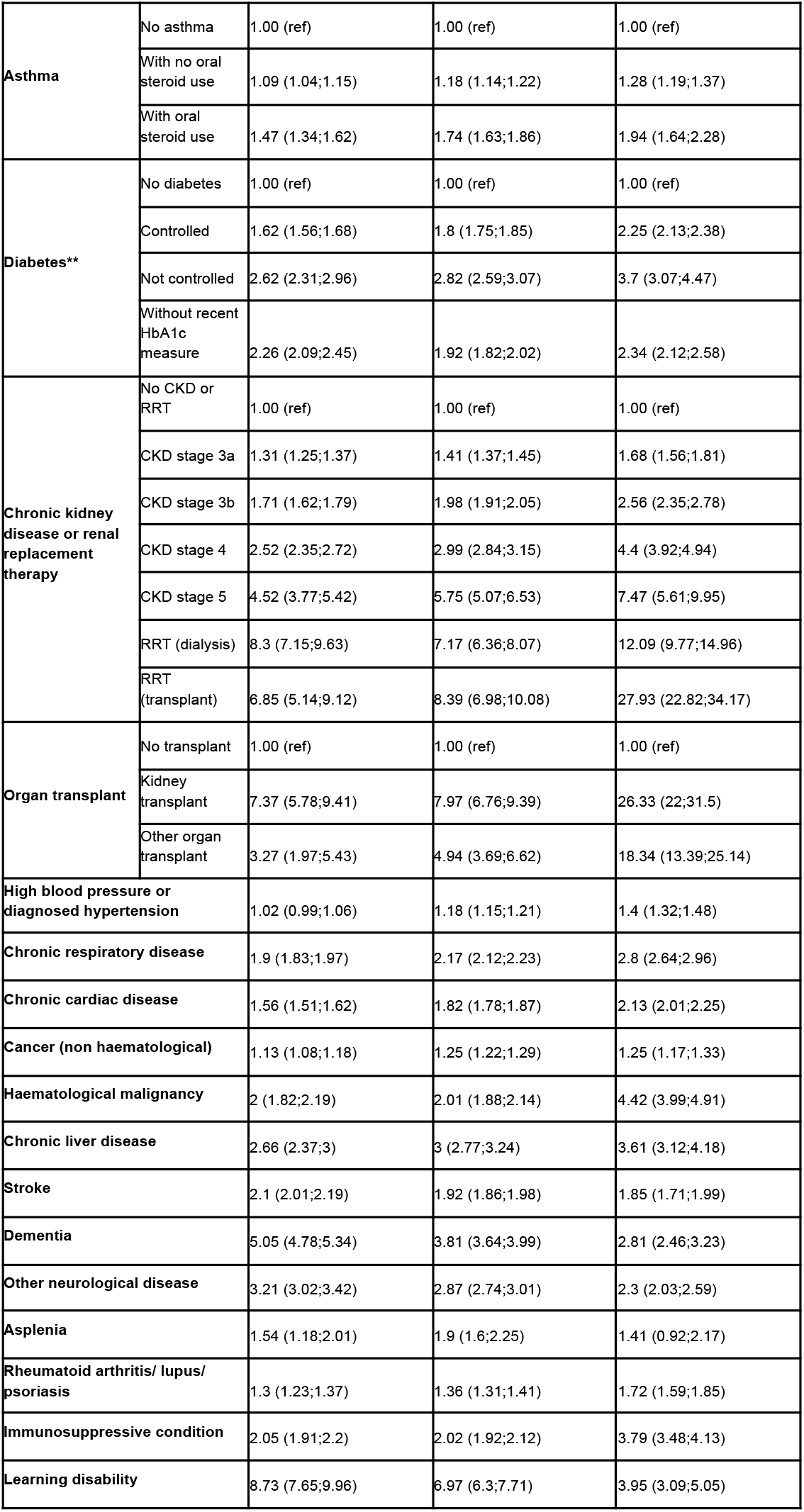
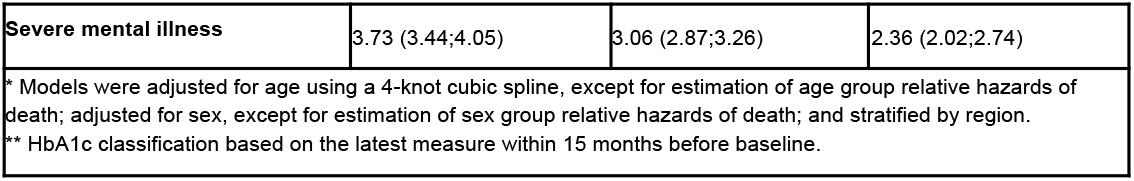
Relative hazard of COVID-19-related death and 95% confidence intervals (95%CI) in OpenSAFELY-TPP in the three pandemic waves in England (wave 1: March 23 to May 30, 2020; wave 2: September 7, 2020 to April 24, 2021; and wave 3, delta: May 28 to December 14, 2021).

## References

[1] E. J. Williamson et al., ‘Factors associated with COVID-19-related death using OpenSAFELY’, Nature, vol. 584, no. 7821, Art. no. 7821, Aug. 2020, doi: 10.1038/s41586-020-2521-4.

[2] M. Woodward, S. A. E. Peters, and K. Harris, ‘Social deprivation as a risk factor for COVID-19 mortality among women and men in the UK Biobank: nature of risk and context suggests that social interventions are essential to mitigate the effects of future pandemics’, J Epidemiol Community Health, vol. 75, no. 11, pp. 1050–1055, Nov. 2021, doi: 10.1136/jech-2020-215810.

[3] R. Mathur et al., ‘Ethnic differences in SARS-CoV-2 infection and COVID-19-related hospitalisation, intensive care unit admission, and death in 17 million adults in England: an observational cohort study using the OpenSAFELY platform’, The Lancet, vol. 397, no. 10286, pp. 1711–1724, May 2021, doi: 10.1016/S0140-6736(21)00634-6.

[4] V. Raleigh, ‘Deaths from Covid-19 (coronavirus)’, The King’s Fund, Apr. 23, 2021. https://www.kingsfund.org.uk/publications/deaths-covid-19 (accessed May 11, 2022).

[5] E. J. Williamson et al., ‘Risks of covid-19 hospital admission and death for people with learning disability: population based cohort study using the OpenSAFELY platform’, BMJ, vol. 374, p. 1592, Jul. 2021, doi: 10.1136/bmj.n1592.

[6] K. Wing et al., ‘Association between household composition and severe COVID-19 outcomes in older people by ethnicity: an observational cohort study using the OpenSAFELY platform’. medRxiv, p. 2022.04.22.22274176, Apr. 22, 2022. doi: 10.1101/2022.04.22.22274176.

[7] ‘Coronavirus (COVID-19) latest insights - Office for National Statistics’. https://www.ons.gov.uk/peoplepopulationandcommunity/healthandsocialcare/conditionsanddiseases/articles/coronaviruscovid19latestinsights/deaths (accessed Mar. 02, 2022).

[8] ‘Impact of COVID-19 vaccines on mortality in England’. https://assets.publishing.service.gov.uk/government/uploads/system/uploads/attachment_data/file/977249/PHE_COVID-19_vaccine_impact_on_mortality_March.pdf (accessed May 11, 2022).

[9] J. H. Beigel et al., ‘Remdesivir for the Treatment of Covid-19 — Final Report’, N. Engl. J. Med., vol. 383, no. 19, pp. 1813–1826, Nov. 2020, doi: 10.1056/NEJMoa2007764.

[10] The WHO Rapid Evidence Appraisal for COVID-19 Therapies (REACT) Working Group, ‘Association Between Administration of Systemic Corticosteroids and Mortality Among Critically Ill Patients With COVID-19: A Meta-analysis’, JAMA, vol. 324, no. 13, pp. 1330–1341, Oct. 2020, doi: 10.1001/jama.2020.17023.

[11] N. G. Davies et al., ‘Effects of non-pharmaceutical interventions on COVID-19 cases, deaths, and demand for hospital services in the UK: a modelling study’, Lancet Public Health, vol. 5, no. 7, pp. e375–e385, Jul. 2020, doi: 10.1016/S2468-2667(20)30133-X.

[12] A. Green et al., ‘Describing the population experiencing COVID-19 vaccine breakthrough following second vaccination in England: A cohort study from OpenSAFELY’. medRxiv, p. 2021.11.08.21265380, Dec. 16, 2021. doi: 10.1101/2021.11.08.21265380.

[13] ‘The R value and growth rate’, GOV.UK. https://www.gov.uk/guidance/the-r-value-and-growth-rate (accessed Mar. 02, 2022).

[14] R. Paton, C. Overton, and T. Ward, ‘The spread of Omicron and replacement of Delta in the UK’. https://assets.publishing.service.gov.uk/government/uploads/system/uploads/attachment_data/file/1046486/S1481_UKHSA_Omicron_growth_advantage.pdf (accessed Mar. 02, 2022).

[15] ‘Who is at high risk from coronavirus (COVID-19)’, Mar. 01, 2021. https://www.nhs.uk/conditions/coronavirus-covid-19/people-at-higher-risk/who-is-at-high-risk-from-coronavirus/ (accessed Mar. 02, 2022).

[16] van Belle, Gerald Fisher, Lloyd D, Heagerty Patrick J., and Lumley Thomas S, ‘Rates and Proportions’, in Biostatistics: A Methodology for the Health Sciences, Second ed., Hoboken, New Jersey, U.S.: John Wiley & Sons, pp. 640–660.

[17] A. R. Y. B. Lee et al., ‘Efficacy of covid-19 vaccines in immunocompromised patients: systematic review and meta-analysis’, BMJ, vol. 376, p. e068632, Mar. 2022, doi: 10.1136/bmj-2021-068632.

[18] E. P. K. Parker et al., ‘Response to additional COVID-19 vaccine doses in people who are immunocompromised: a rapid review’, Lancet Glob. Health, vol. 10, no. 3, pp. e326–e328, Mar. 2022, doi: 10.1016/S2214-109X(21)00593-3.

[19] ‘COVID-19: the green book, chapter 14a’, GOV.UK. https://www.gov.uk/government/publications/covid-19-the-green-book-chapter-14a (accessed Jul. 21, 2022).

[20] H. J. Curtis et al., ‘Trends and clinical characteristics of COVID-19 vaccine recipients: a federated analysis of 57.9 million patients’ primary care records in situ using OpenSAFELY’, Br. J. Gen. Pract., vol. 72, no. 714, pp. e51–e62, Jan. 2022, doi: 10.3399/BJGP.2021.0376.

[21] S. Mouffak, Q. Shubbar, E. Saleh, and R. El-Awady, ‘Recent advances in management of COVID-19: A review’, Biomed. Pharmacother., vol. 143, p. 112107, Nov. 2021, doi: 10.1016/j.biopha.2021.112107.

[22] ‘Hospitalized Adults: Therapeutic Management’, COVID-19 Treatment Guidelines. https://www.covid19treatmentguidelines.nih.gov/management/clinical-management/hospitalized-adults--therapeutic-management/ (accessed Jul. 21, 2022).

[23] ‘Timeline of UK government coronavirus lockdowns and restrictions’, The Institute for Government. https://www.instituteforgovernment.org.uk/charts/uk-government-coronavirus-lockdowns (accessed Jul. 25, 2022).

[24] S. Talic et al., ‘Effectiveness of public health measures in reducing the incidence of covid-19, SARS-CoV-2 transmission, and covid-19 mortality: systematic review and meta-analysis’, BMJ, vol. 375, p. e068302, Nov. 2021, doi: 10.1136/bmj-2021-068302.

[25] W. S. Hart et al., ‘Generation time of the alpha and delta SARS-CoV-2 variants: an epidemiological analysis’, Lancet Infect. Dis., vol. 22, no. 5, pp. 603–610, May 2022, doi: 10.1016/S1473-3099(22)00001-9.

[26] C. Andrews et al., ‘OpenSAFELY: Representativeness of electronic health record platform OpenSAFELY-TPP data compared to the population of England’. Wellcome Open Research, Jul. 18, 2022. doi: 10.12688/wellcomeopenres.18010.1.

[27] W. J. Hulme et al., ‘Comparative effectiveness of ChAdOx1 versus BNT162b2 covid-19 vaccines in health and social care workers in England: cohort study using OpenSAFELY’, BMJ, vol. 378, p. e068946, Jul. 2022, doi: 10.1136/bmj-2021-068946.

[28] S. J. W. Evans and N. P. Jewell, ‘Vaccine Effectiveness Studies in the Field’, N. Engl. J. Med., vol. 385, no. 7, pp. 650–651, Aug. 2021, doi: 10.1056/NEJMe2110605.

[29] ‘Interim recommendations for an extended primary series with an additional vaccine dose for COVID-19 vaccination in immunocompromised persons’. https://www.who.int/publications-detail-redirect/WHO-2019-nCoV-vaccines-SAGE_recommendation-immunocompromised-persons (accessed Jul. 25, 2022).

[30] ‘Data Security and Protection Toolkit’, NHS Digital. https://digital.nhs.uk/data-and-information/looking-after-information/data-security-and-information-governance/data-security-and-protection-toolkit (accessed Jul. 26, 2022).

[31] ‘ISB1523: Anonymisation Standard for Publishing Health and Social Care Data’, NHS Digital. https://digital.nhs.uk/data-and-information/information-standards/information-standards-and-data-collections-including-extractions/publications-and-notifications/standards-and-collections/isb1523-anonymisation-standard-for-publishing-health-and-social-care-data (accessed Jul. 26, 2022).

[32] ‘Coronavirus (COVID-19): notification to organisations to share information - GOV.UK’. https://web.archive.org/web/20200421171727/https://www.gov.uk/government/publications/coronavirus-covid-19-notification-of-data-controllers-to-share-information (accessed Jul. 26, 2022).

[33] ‘Coronavirus (COVID-19): notice under Regulation 3(4) of the Health Service (Control of Patient Information) Regulations 2002’, GOV.UK. https://www.gov.uk/government/publications/coronavirus-covid-19-notification-to-organisations-to-share-information/coronavirus-covid-19-notice-under-regulation-34-of-the-health-service-control-of-patient-information-regulations-2002 (accessed Jul. 26, 2022).

[34] ‘Confidentiality Advisory Group’, Health Research Authority. https://www.hra.nhs.uk/about-us/committees-and-services/confidentiality-advisory-group/ (accessed Jul. 26, 2022).

